# Association of lipid-regulating drugs with dementia and related conditions: an observational study of data from the Clinical Practice Research Datalink

**DOI:** 10.1101/2021.10.21.21265131

**Authors:** Luke A McGuinness, Julian PT Higgins, Venexia M Walker, Neil M Davies, Richard M Martin, Elizabeth Coulthard, George Davey-Smith, Patrick G Kehoe, Yoav Ben-Shlomo

## Abstract

2

**Background:** There is some evidence that circulating blood lipids play a role in the development of Alzheimer’s disease (AD) and dementia. These modifiable risk factors could be targeted by existing lipid-regulating agents, including statins, for dementia prevention. Here, we test the association between lipid-regulating agents and incidence of dementia and related conditions in the Clinical Practice Research Datalink (CPRD), an United Kingdom-based electronic health record database.

**Methods:** A retrospective cohort study was performed using routinely collected CPRD data (January 1995 and March 2016). Multivariable Cox proportional hazard models, allowing for a time-varying treatment indicator, were used to estimate the association between seven lipid-regulating drug classes (vs. no drug) and five dementia outcomes (all-cause, vascular and other dementias, and probable and possible Alzheimer’s disease).

**Results:** We analyzed 1,684,564 participants with a total follow-up of 10,835,685 patient-years (median: 5.9 years (IQR:2.7-9.7)). We found little evidence that lipid-regulating agents were associated with incidence of Alzheimer’s disease (probable HR:0.98, 95%CI:0.94-1.01; possible HR:0.97, 95%CI:0.93-1.01), but there was evidence of an increased risk of all-cause (HR:1.17, 95%CI:1.14-1.19), vascular (HR:1.81, 95%CI:1.73-1.89) and other dementias (HR:1.19, 95%CI:1.15-1.24). Evidence from a number of control outcomes indicated the presence of substantial residual confounding by indication (ischaemic heart disease HR: 1.62, 95%CI: 1.59-1.64; backpain HR: 1.04, 95%CI: 1.03-1.05; and Type 2 diabetes HR: 1.50, 95%CI: 1.48-1.51).

**Conclusion:** Lipid-regulating agents were not associated with reduced Alzheimer’s disease risk. There was some evidence of an increased the risk of all-cause, vascular and other dementias, likely due to residual confounding by indication.

**Key messages:**

- A large cohort of patients from the Clinical Practice Research Datalink (CPRD) electronic health record database was assembled to examine the association of lipid-regulating agents, such as statins, with dementia outcomes.
- There was little evidence that lipid-regulating agents were associated with Alzheimer’s disease, but there was some evidence for a harmful association with all-cause, vascular and other dementias. In all cases, the estimated associations were driven by the any statin subgroup, which comprised most participants in our cohort.
- Evidence from the control outcome analyses indicated strong residual confounding by indication, mostly likely related to vascular factors.

## 4 Introduction

Dementia is a major progressive neurocognitive disorder, the most common types of which are Alzheimer’s disease, vascular dementia and Lewy Body dementia.(1) Despite an increasing number of cases globally and decades of research, there remains much unknown about the pathogenesis and progression of the disease, and, at present, no effective treatment exists to arrest, slow or reverse the cognitive decline associated with the condition.(2) Drug repurposing, the identification of new applications for previously approved drugs, may provide an efficient mechanism to discover new effective preventative and therapeutic treatments for dementia.(3,4)

Several cardiovascular factors have been identified as potential risk factors for dementia,(5) and of these, circulating lipid levels represent a promising target for intervention due to the ready availability of lipid-modifying treatments. In this context, determining whether lipid-regulating agents (LRA) could be repurposed for the prevention of dementia and related diseases would be helpful in the development of evidence-based prevention policy. Several existing prospective studies have examined the association of LRA use with dementia.(6–10) However, many of these studies are small, record few outcomes, and had limited follow-up.

The use of electronic health data for epidemiological research has several advantages.(11) As the data are collected through the routine care of a large cohort, they allow for nested cohort studies using sample sizes and time-scales which would be infeasible using traditional methods. In addition, data are collected for care provision and without a specific research question in mind, providing a holistic picture of a patient and their health experience. While they may not contain information on all potentially important confounders,(12) electronic health records provide routine data on common covariates for millions of participants.

We therefore aimed to test the association between several major classes of LRA and all-cause dementia, Alzheimer’s disease, vascular dementia and other dementias, in the Clinical Practice Research Datalink (CPRD), a large, population-based electronic health record (EHR) database from the United Kingdom.(13)

## 5 Methods

### 5.1 Study design and protocol

We defined a longitudinal cohort using data from the CPRD. Our initial sample included all participants registered at a participating practice between 1 January 1995 and 29 February 2016 who had a flag for “research quality” data. Records pre-dating the 1995 cut-off were excluded from the analysis as data quality and reliability are thought to be higher after this date.(14) All events of interest were identified using predetermined code lists, which are available for inspection (see Data/code availability).

An *a priori* protocol for this study was published,(15) and amendments to this are recorded in Supplementary Materials 1. This study was reported in line with the RECORD guidelines (Supplementary Materials 2).(16)

### 5.2 Study Cohort

Participants were included in our study cohort if their record contained any of the following index events: (a) a code for a diagnosis of hypercholesterolemia or related condition; (b) a code for prescription of a LRA (such as statins); (c) a total cholesterol test result of >4mmol/L; (d) or a low-density lipoprotein cholesterol (LDL-c) test result of >2mmol/L.

These index events allowed us to define a population of participants who were either at risk of hypercholesterolemia, as indicated by the elevated total or LDL-c test results, or had already been diagnosed with it, as indicated by a diagnostic code or related prescription. This approach, conditioning entry into the study on being either “at-risk” or already diagnosed with hypercholesterolemia, was employed to reduce the “confounding by indication” that we would expect to observe if we had used a general population cohort.

An index date for each participant was defined as the date when the first relevant code or test result (as detailed above) was recorded on their clinical record. Participants were followed up until the earliest of (a) an outcome of interest; (b) death; (c) end of follow-up (29 February 2016); (d) last registration date with their GP practice; or (e) the last CPRD collection date for their practice. Participants were removed from our sample if they were less than 40 years of age at entry (as these patients are less likely to be prescribed a LRA), had less than 12 months of “research quality” data prior to their index date, were simultaneously prescribed more than one LRA (due to the difficulty of assigning these patients to a single exposure group), or were diagnosed with dementia before or on the date of the index event.

### 5.3 Exposures

We considered seven lipid-regulating drug classes based on groupings in the British National Formulary (BNF)(17), namely: statins, fibrates, bile acid sequestrants, ezetimibe, nicotinic acid groups, ezetimibe and statin (representing one treatment containing both drugs, rather than the two classes being prescribed concurrently), and omega-3 fatty acid groups.

To address the potential for immortal time bias, we employed a time-varying indicator of treatment status to correctly allocate time-at-risk to the exposed and unexposed groups.(18) Under this model, all participants entered the unexposed group on their index date and were moved into the exposed group on the date they were first prescribed an eligible LRA. Participants whose index event was a LRA prescription entered the study and the exposed group on the same day, and so contributed no time-at-risk to the unexposed group.

A participant’s drug class was assigned based on their first recorded prescription, and any drug switching was ignored to mimic an intention-to-treat approach. We did however tabulate how often the initial drug class was stopped (defined as last prescription of the primary class being followed by at least six months of observation), added to (defined as a second drug class being prescribed before the last prescription of the initial class), or switched (defined as a second drug class being prescribed after the last prescription of the initial class).

### 5.4 Outcomes

We considered five outcomes as part of this analysis: probable Alzheimer’s disease, possible Alzheimer’s disease, vascular dementia, other dementias, and a composite all-cause dementia outcome. When two or more outcomes were coded in a participant’s clinical record, a decision tree was used to differentiate between them (Supplementary Figure 1). The diagnosis date of the outcome was determined by the first record of a relevant code.

### 5.5 Covariates

The analysis was adjusted for a range of baseline covariates including sex, grouped year of entry into the cohort (<=2000, 2001-2005, 2006-2010, >2010), Charlson co-morbidity index, Index of Multiple Deprivation (IMD), consultation rate, alcohol use (current, former, never), smoking (current, former, never), BMI, baseline total cholesterol, and history of cardiovascular disease, coronary bypass surgery, coronary artery disease, peripheral arterial disease, hypertension, chronic kidney disease, and Type 1 and Type 2 diabetes. These variables were selected as potential confounders between dementia outcomes and use of a LRA. All covariates were determined at the index date and definitions for each can be found in Supplementary Table 1.

### 5.6 Analysis plan

All analyses were performed in STATA (V16, StataCorp LLC, College Station, TX). Cox proportional hazard models with a time-varying treatment indicator were used to estimate the hazard ratio and corresponding 95% confidence intervals, allowing for potential clustering of outcomes by practice. Participant’s age was used as the time axis for all models.(19–21) To observe the effect of adjusting for additional covariates, we compared models adjusted for age only and age and sex with the fully adjusted model. Additional analyses stratified by outcome and drug class were also performed.

In the case of missing data, we used multiple imputation by chained equations (MICE) in STATA to create 20 imputed datasets.(22) All covariates included in the analytic model were also included in the imputation model.(23) The full imputation model is available for inspection (See Data/Code availability section).

### 5.7 Sensitivity analyses

We performed several sensitivity analyses. As statins are contraindicated in pregnancy,(24) we ran the models described above but excluding participants below the age of 55. Given the different ability of lipophilic statins to cross the blood-brain barrier,(25) we further stratified the statin exposure group into lipophilic (Atorvastatin, Lovastatin, Simvastatin, Cerivastatin) and hydrophilic (Pravastatin, Rosuvastatin, Fluvastatin) statins. Finally, we included three alternative outcomes with known associations with statin use as positive or negative controls using the fully adjusted model: back pain (negative control), ischaemic heart disease (positive protective control), and Type 2 diabetes (positive harmful control).(26,27)

## 6 Results

### 6.1 Patient characteristics

A total of 1,684,564 participants met the inclusion criteria for our cohort (see Supplementary Figure 2 for the attrition flowchart), with a total follow-up of 10,835,685 patient years at risk. Most participants were included in the cohort due to an elevated test result (elevated cholesterol test result: 93%, prescription of LRA: 5.6%, code for hypercholesterolemia: 1%). The median age at the index date was 57 years (inter-quartile range (IQR):48-67) and participants were followed up for a median of 5.9 years (IQR:2.7-9.7). During follow-up, an all-cause dementia diagnosis was recorded for 41,830 patients (12,647 probable Alzheimer’s disease, 9,954 possible Alzheimer’s disease, 8,466 vascular dementia, 10,763 other dementia).

The number of events, time-at-risk and crude rates for each drug class, tabulated by dementia outcome, are shown in Table 1. Most participants (98.1%) prescribed a lipid-regulating agent were prescribed a statin. We excluded the “Ezetimibe and statins” and “Nicotinic acid groups” drug classes from subsequent subgroup analyses based on the extremely small number of participants in these groups (Table 1). The distribution of baseline characteristics across the remaining seven drug classes can be seen in Table 2. The stopping, addition and switching of drug classes was common across all exposure groups (Supplementary Table 2).

**Table 1:** Summary of number of events, participant-time-at-risk and crude rates by drug class and dementia outcome.

| Exposure group | Any dementia |  |  | Probable AD |  |  | Possible AD |  |  | Vascular dementia |  |  | Other dementia |  |  |
| --- | --- | --- | --- | --- | --- | --- | --- | --- | --- | --- | --- | --- | --- | --- | --- |
|  | Events | PYAR | Rate<sup>a</sup> | Events | PYAR | Rate<sup>a</sup> | Events | PYAR | Rate<sup>a</sup> | Events | PYAR | Rate<sup>a</sup> | Events | PYAR | Rate<sup>a</sup> |
| No LRA (unexposed) | 18,608 | 5,872,717 | 317 | 6,368 | 5,818,047 | 109 | 2,637 | 5,800,964 | 45 | 4,813 | 5,811,594 | 83 | 4,790 | 5,808,285 | 82 |
| By drug class |  |  |  |  |  |  |  |  |  |  |  |  |  |  |  |
| Statins | 22,920 | 4,871,568 | 470 | 6,190 | 4,758,526 | 130 | 5,773 | 4,753,437 | 121 | 5,871 | 4,755,258 | 123 | 5,086 | 4,747,237 | 107 |
| Omega-3 FAGs | 19 | 8,034 | 236 | 4 | 7,927 | 50 | 7 | 7,950 | 88 | 4 | 7,938 | 50 | 4 | 7,925 | 50 |
| Fibrates | 141 | 38,003 | 371 | 49 | 37,102 | 132 | 21 | 36,835 | 57 | 36 | 37,001 | 97 | 35 | 36,983 | 95 |
| Ezetimibe | 32 | 6,604 | 485 | 8 | 6,429 | 124 | 7 | 6,425 | 109 | 12 | 6,444 | 186 | 5 | 6,393 | 78 |
| BAS | 106 | 36,370 | 291 | 28 | 35,808 | 78 | 19 | 35,726 | 53 | 26 | 35,768 | 73 | 33 | 35,808 | 92 |
| Ezetimibe + Statins<sup>b</sup> | 0 | 986 | - | 0 | 986 | - | 0 | 986 | - | 0 | 986 | - | 0 | 986 | - |
| NAG | 4 | 1,403 | - | 0 | 1,379 | - | 2 | 1,391 | - | 1 | 1,389 | - | 1 | 1,382 | - |
| Total | 41,830 | 10,835,686 | 386 | 12,647 | 10,666,205 | 119 | 8,466 | 10,643,714 | 80 | 10,763 | 10,656,378 | 101 | 9,954 | 10,644,999 | 94 |
| <sup>a</sup> Crude rate per 100,000 participant-years-at-risk<br><sup>b</sup> One treatment containing both drugs, rather than the two classes being prescribed concurrently<br>Abbreviations: AD - Alzheimer's disease; BAS - Bile acid sequestrants; LRA - Lipid regulating agent; NAG - Nicotinic acid groups; Omega-3 FAGs - Omega-3 fatty acid groups; PYAR - Participant-years-at-risk. |  |  |  |  |  |  |  |  |  |  |  |  |  |  |  |

**Table 2:**
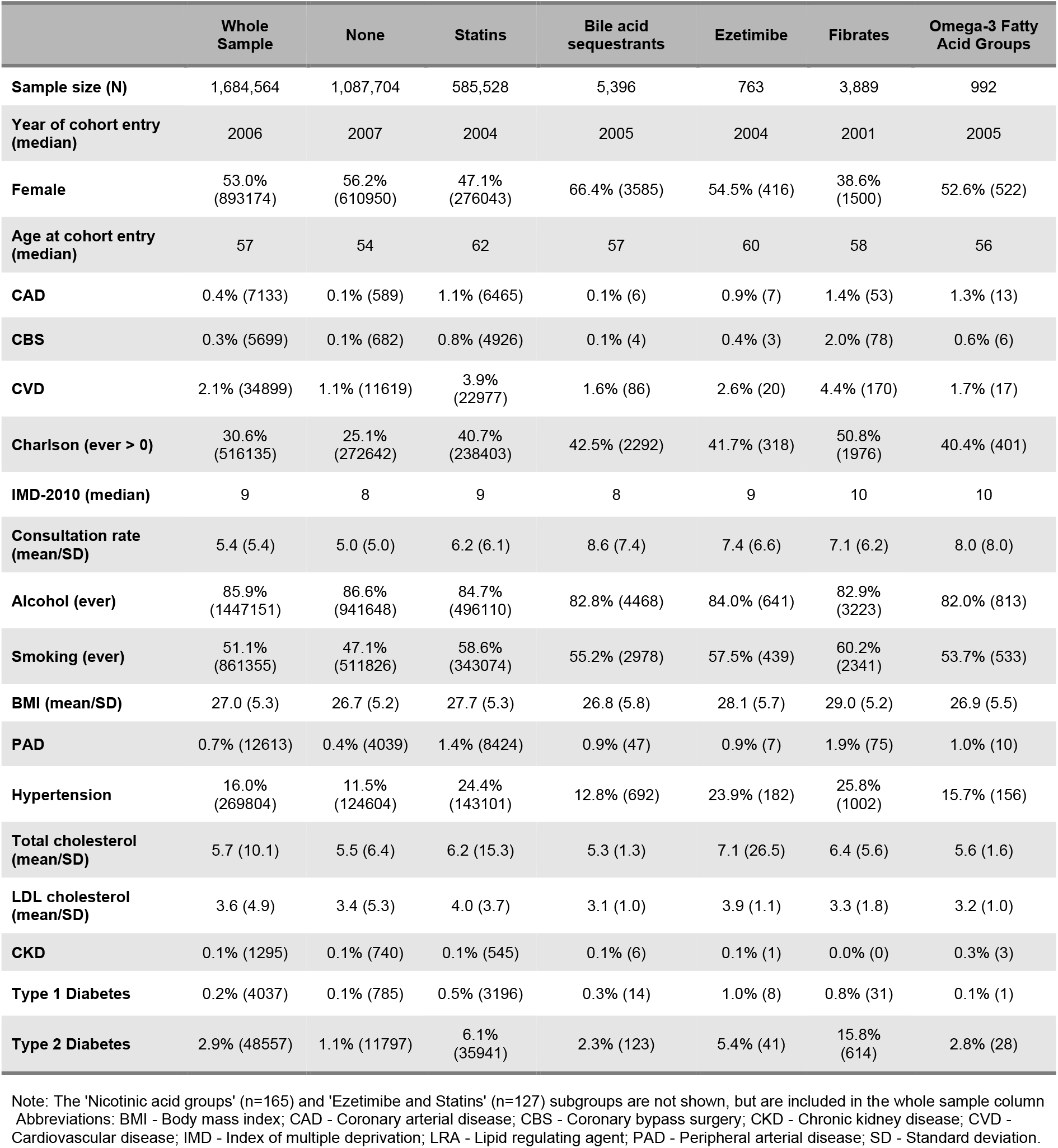
Patient characteristics by drug class. Summary statistics are presented as “% (N)” unless otherwise specified in the variable name.

|  | Whole Sample | None | Statins | Bile acid sequestrants | Ezetimibe | Fibrates | Omega-3 Fatty Acid Groups |
| --- | --- | --- | --- | --- | --- | --- | --- |
| **Sample size (N)** | 1,684,564 | 1,087,704 | 585,528 | 5,396 | 763 | 3,889 | 992 |
| **Year of cohort entry (median)** | 2006 | 2007 | 2004 | 2005 | 2004 | 2001 | 2005 |
| **Female** | 53.0% (893174) | 56.2% (610950) | 47.1% (276043) | 66.4% (3585) | 54.5% (416) | 38.6% (1500) | 52.6% (522) |
| **Age at cohort entry (median)** | 57 | 54 | 62 | 57 | 60 | 58 | 56 |
| **CAD** | 0.4% (7133) | 0.1% (589) | 1.1% (6465) | 0.1% (6) | 0.9% (7) | 1.4% (53) | 1.3% (13) |
| **CBS** | 0.3% (5699) | 0.1% (682) | 0.8% (4926) | 0.1% (4) | 0.4% (3) | 2.0% (78) | 0.6% (6) |
| **CVD** | 2.1% (34899) | 1.1% (11619) | 3.9% (22977) | 1.6% (86) | 2.6% (20) | 4.4% (170) | 1.7% (17) |
| **Charlson (ever > 0)** | 30.6% (516135) | 25.1% (272642) | 40.7% (238403) | 42.5% (2292) | 41.7% (318) | 50.8% (1976) | 40.4% (401) |
| **IMD-2010 (median)** | 9 | 8 | 9 | 8 | 9 | 10 | 10 |
| **Consultation rate (mean/SD)** | 5.4 (5.4) | 5.0 (5.0) | 6.2 (6.1) | 8.6 (7.4) | 7.4 (6.6) | 7.1 (6.2) | 8.0 (8.0) |
| **Alcohol (ever)** | 85.9% (1447151) | 86.6% (941648) | 84.7% (496110) | 82.8% (4468) | 84.0% (641) | 82.9% (3223) | 82.0% (813) |
| **Smoking (ever)** | 51.1% (861355) | 47.1% (511826) | 58.6% (343074) | 55.2% (2978) | 57.5% (439) | 60.2% (2341) | 53.7% (533) |
| **BMI (mean/SD)** | 27.0 (5.3) | 26.7 (5.2) | 27.7 (5.3) | 26.8 (5.8) | 28.1 (5.7) | 29.0 (5.2) | 26.9 (5.5) |
| **PAD** | 0.7% (12613) | 0.4% (4039) | 1.4% (8424) | 0.9% (47) | 0.9% (7) | 1.9% (75) | 1.0% (10) |
| **Hypertension** | 16.0% (269804) | 11.5% (124604) | 24.4% (143101) | 12.8% (692) | 23.9% (182) | 25.8% (1002) | 15.7% (156) |
| **Total cholesterol (mean/SD)** | 5.7 (10.1) | 5.5 (6.4) | 6.2 (15.3) | 5.3 (1.3) | 7.1 (26.5) | 6.4 (5.6) | 5.6 (1.6) |
| **LDL cholesterol (mean/SD)** | 3.6 (4.9) | 3.4 (5.3) | 4.0 (3.7) | 3.1 (1.0) | 3.9 (1.1) | 3.3 (1.8) | 3.2 (1.0) |
| **CKD** | 0.1% (1295) | 0.1% (740) | 0.1% (545) | 0.1% (6) | 0.1% (1) | 0.0% (0) | 0.3% (3) |
| **Type 1 Diabetes** | 0.2% (4037) | 0.1% (785) | 0.5% (3196) | 0.3% (14) | 1.0% (8) | 0.8% (31) | 0.1% (1) |
| **Type 2 Diabetes** | 2.9% (48557) | 1.1% (11797) | 6.1% (35941) | 2.3% (123) | 5.4% (41) | 15.8% (614) | 2.8% (28) |

Note: The 'Nicotinic acid groups' (n=165) and 'Ezetimibe and Statins' (n=127) subgroups are not shown, but are included in the whole sample column Abbreviations: BMI - Body mass index; CAD - Coronary arterial disease; CBS - Coronary bypass surgery; CKD - Chronic kidney disease; CVD - Cardiovascular disease; IMD - Index of multiple deprivation; LRA - Lipid regulating agent; PAD - Peripheral arterial disease; SD - Standard deviation.

### 6.2 Missing data

Full covariate information was available for 450,234 participants (26.7%). Five key variables had some missing data: IMD 2010 score, a proxy for socioeconomic position that is measured as twentiles with 1 indicating the least deprived and 20 indicating the most deprived, was missing for 625,788 participants (37.1%), because it is only recorded for selected English practices; alcohol status was missing for 269,526 participants (16%); smoking status was missing for 84,424 participants (5%); BMI, or a calculated BMI from height and weight measurements, was missing for 266,672 participants (15.8%); baseline total cholesterol was missing for 119,675 participants (7.1%); and baseline LDL cholesterol was missing for 787,289 participants (46.7%).

### 6.3 Primary analysis

#### Alzheimer’s disease

As shown in Figure 1, our results show little evidence of an association between lipid-regulating agents and probable (HR:0.98, 95%CI:0.94-1.01) or possible (HR:0.97, 95%CI:0.93-1.01) Alzheimer’s disease when compared with no treatment, except for an adverse association between fibrates and probable Alzheimer’s disease (HR:1.52, 95%CI:1.13-2.03).

**Figure 1:**
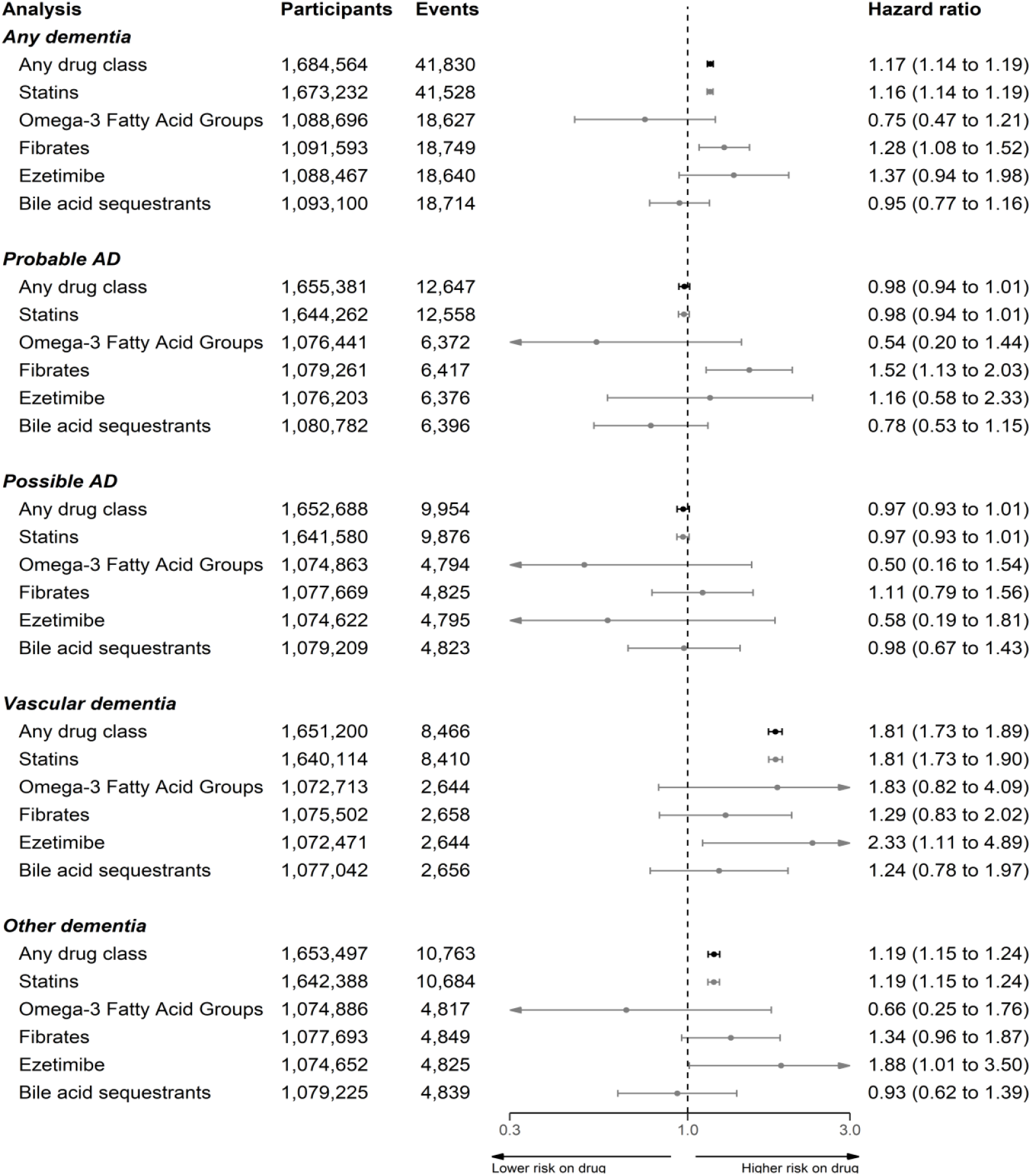
Results from the primary analyses of CPRD data comparing prescription of an lipid-regulating drug with no prescription, stratified by dementia outcome and drug class. All results were obtained using the fully adjusted model and participant age as the time scale.

#### Non-Alzheimer’s disease dementias

In contrast to the findings for Alzheimer’s disease outcomes, lipid-regulating agents were associated with an increased risk of a subsequent diagnosis of vascular dementia (HR:1.81, 95%CI:1.73-1.89) or other dementias (HR:1.19, 95%CI:1.15-1.24). Again, the observed estimate was driven mainly by the any statin subgroup, but there was some evidence that ezetimibe was associated with an increased risk of vascular (HR:2.33, 95%CI:1.11-4.89) and other (HR:1.88, 95%CI:1.01-3.5) dementias.

#### All-cause dementia

For the composite all-cause dementia outcome, we found treatment with a lipid-regulating agent was associated with a slightly increased risk (HR:1.17, 95%CI:1.14-1.19), which lies between the associations for the Alzheimer and non-Alzheimer dementia outcomes as would be expected. There was also some evidence that fibrates were associated with increased risk of all-cause dementia (HR:1.28, 95%CI:1.08-1.52).

### 6.4 Sensitivity analyses

Adjustment for additional covariates beyond age and sex had a limited impact (Supplementary Figure 3), except for the probable Alzheimer’s disease outcome, where the full adjustment attenuated to the null the protective association observed when adjusting only for age and sex.

Removing participants aged 55 and under at the index date from our analysis had minimal effect on our estimates (Supplementary Figure 4). When stratifying by statin properties, hydrophilic statins were less harmful in relation to the any, vascular and other dementias outcomes compared with lipophilic statins (Supplementary Figure 5). Similarly, hydrophilic statins were associated with a reduced incidence of Alzheimer’s disease, compared with the absence of evidence for an association with lipophilic statins.

For our control outcomes (Supplementary Figure 6), there was some evidence that patients prescribed a lipid-regulating agent had an increased risk of back pain (HR: 1.04, 95%CI: 1.03-1.05), ischaemic heart disease (HR: 1.62, 95%CI: 1.59-1.64) and Type 2 diabetes (HR: 1.50, 95%CI: 1.48-1.51).

## 7 Discussion

### 7.1 Main findings

There was little evidence for an association between lipid-regulating agents and probable and possible Alzheimer’s when compared with no treatment, but some evidence they were associated with an increased risk of an all-cause dementia, secondary to their association with vascular and other dementias diagnoses. The association observed in each case was driven by the any statin subgroup, which included a substantial majority of participants. For the other drug classes, there was limited evidence of an association with any outcome, with two exceptions. Ezetimibe was associated with increased risk of vascular and other dementias, while fibrates were associated with increased risk of all-cause dementia and probable Alzheimer’s disease.

### 7.2 Comparison with other literature

Much of the existing literature focuses on the association of statins alone with neurodegenerative outcomes, with other lipid-regulating agents being grouped as “non-statin cholesterol-lowering drugs,”(8) echoing the distribution of participants in our analysis.

#### Statins and all-cause dementia

A recent Cochrane Review identified two randomized trials comparing treatment with statins versus non-treatment for the prevention of dementia, only one of which presented information on the incidence of dementia.(28) This study (Heart Protection Study) showed no effect of treatment with simvastatin on all-cause dementia risk (OR: 1.00, 95%CI:0.61-1.65),(29) but concerns were raised over the diagnostic criteria used. A meta-analysis of 30 observational studies found a reduced risk of all-cause dementia was associated with statin treatment (RR 0.83, 95%CI: 0.79–0.87).(30) This conflicts with the findings of our analysis, where statin use was associated with an increased risk of all-cause dementia (HR:1.17, 95%CI:1.14-1.19). Some of the included studies in the meta-analysis specifically exclude vascular dementia from the definition of all-cause dementia,(31) which may limit the ability for comparison with our findings for the all-cause dementia outcome.

Additionally, a previous analysis of the THIN EHR database using a propensity-score matched analysis found a protective effect of statins on all-cause dementia (HR:0.81, 95%CI:0.69-0.96)(32). Differences in the code-lists used to define dementia outcomes, in addition an analytical approach that adjusted for covariates defined after the index date, may go some way to explaining the discrepancy.

#### Statins and Alzheimer’s disease

Our results are broadly in line with the findings of two distinct approaches examining the effect of statin treatment on subsequent Alzheimer’s disease. No randomized trials of statins for the prevention of Alzheimer’s disease have been reported, but a recent meta-analysis of 20 observational studies found statins were associated with a reduced risk of Alzheimer’s disease (RR 0.69, 95% CI 0.60–0.80) with stronger evidence than observed in our analysis.(30) This review included case-control studies and analyses likely to be at risk of immortal time bias, which may account for the discrepancy with our findings. Additionally, a recent Mendelian randomization study examining the effect of genetic inhibition of HMGCR on Alzheimer’s disease (a genetic proxy for statin treatment) provided equivocal evidence (OR: 0.91, 95%CI: 0.63-1.31) but was consistent with our results.(33)

Our additional analyses stratified by statin properties found little evidence of differences in associations of lipophilic and hydrophilic statins and incidence of Alzheimer’s disease, consistent with a recent meta-analysis of observational studies.(6)

#### Statins and vascular/other dementia

Far fewer studies have tested the association between lipid-regulating agents and vascular dementia or other dementias. A recent review found four observational studies examining the association of statins and vascular dementia found limited evidence for an effect (RR:0.93, 95% CI 0.74–1.16).(30) This contrasts with the harmful association found in our analysis (HR:1.81, 95%CI:1.73-1.89). When stratifying by lipid properties, lipophilic statins were more harmful than hydrophilic statins in vascular dementia, potentially due to their ability to cross the blood brain barrier.

#### Other drug classes

Apart from statins, few studies examining a lipid-regulating agent have been reported. One of the few classes for which a previous analysis was available were fibrates, which found little evidence of an association with all-cause dementia was identified,(8) inconsistent with our finding that patients prescribed fibrates had higher all-cause dementia risk than those prescribed other lipid lowering agents.

A previous Mendelian randomization study found little evidence that genetic variants that proxy for ezetimibe affect risk of Alzheimer’s disease (OR: 1.17, 95%CI: 0.73-1.87),(33) consistent with our findings. To our knowledge, there is no previous study of the effect of preventative treatment with ezetimibe on risk of vascular dementia.

### 7.3 Strengths and limitations

A major strength of our analysis is the size of the included cohort and the length of follow-up that the use of electronic health records allowed. In addition, we followed users and non-users from a common index date, using a time-updating treatment indicator to correctly assign time-at-risk to the exposed and unexposed groups.

However, the findings of our analysis are subject to several limitations. Confounding by indication is a major bias in pharmacoepidemiological studies and could provide a potential explanation for the observed increased risk of vascular and other dementias with lipid regulating agent use. Patients with vascular risk factors are more likely to receive a statin prescription and also to be diagnosed with vascular dementia. Supporting evidence for this interpretation comes from a variety of sources, including the results of the control outcome analyses. This would explain the increased risk for the ischaemic heart disease outcome, for which statins are known to be protective, whilst almost no association was observed with the backpain outcome, indicating that most of the uncontrolled confounding is likely to be related to vascular factors. Additionally, we obtained the expected harmful result for Type 2 diabetes, where statins’ mechanism of action on this outcome is unlikely to be vascular.(26,34,35) Further supporting evidence comes from the increasingly harmful association observed when moving from the probable and possible Alzheimer’s disease outcome to the other dementias outcome, and finally to the vascular dementia outcomes. This pattern suggests that the strength of the residual confounding by indication increases as the proportion of cases with a vascular component in an outcome definition increases. A review of other available literature suggests that this observation (a harmful effect of lipid regulating agents on vascular-related outcome due to confounding by indication) is not unusual. Using a conventional epidemiological technique, a previous analysis also found an increased risk of coronary heart disease (analogous to the ischaemic heart disease outcome used in our analysis) in those taking statins (HR: 1.31, 95%CI: 1.04-1.66).(36) Controlling for confounding by indication in that study through the use of a trial emulation analysis gave an estimate of 0.89 (95%CI: 0.73-1.09), a comparable though less precise estimate to that observed in RCTs of statin use (0.73, 95%CI: 0.67-0.80).(37)

A secondary limitation is the potential for differential outcome misclassification based on the exposure, as we cannot exclude the possibility that for people with memory complaints, a diagnosis of vascular dementia might be made more frequently than Alzheimer’s disease if their medical records contain prescriptions for lipid-regulating agents. Further, there is also the potential for non-differential misclassification of the outcome, based on the use of electronic health records to identify dementia cases.(38,39)

A further limitation stems from uncontrolled confounding due to genetic factors. Number of ApoE *ϵ*4 alleles represents the strongest genetic risk factor for Alzheimer’s disease, but also substantially increases LDL-c levels,(40) potentially prompting treatment with a statin or other lipid regulating agent. We were unable to control for ApoE genotype in this analysis as we did not have access to genetic data on participants. As a result, any protective association between LRA use and the Alzheimer’s disease outcomes may be masked by residual negative confounding by ApoE.

Finally, as with many studies of dementia, there is a risk of reverse causation in our analysis. Dementia and associated conditions have a long prodromal period, during which preclinical disease could cause indications for the prescription of a lipid-regulating agent.

## 8 Conclusions

We have provided new evidence on the association of lipid-regulating agent prescription, predominantly statins, with all-cause dementia, Alzheimer’s disease, vascular dementia, and other dementias. We found limited evidence that use of lipid-regulating agents was associated with incidence of probable or possible Alzheimer’s disease, but they were associated with an increased risk of all-cause, vascular and other dementias. Despite our attempts to account for bias in our analysis, there is a strong potential for residual confounding, misclassification and reverse causation.

This is likely to explain the unexpected increase in risk of vascular dementia associated with statin use. Future research should aim to address these biases using newer methods such as a trial emulation analysis.

## Supporting information

Codelists

## Data Availability

This analysis used the CPRD-GOLD primary care dataset March 2016 snapshot (ISAC 15_246R), which is available upon application to the CPRD Independent Scientific Advisory Committee. The code lists used to define the outcomes and covariates for this study, in addition to the scripts used to create the study cohort and perform the analyses, are available on GitHub (https://github.com/mcguinlu/CPRD-LRA).

## 9 Declarations

### Ethics approval

The protocol for this study was approved by the CPRD’s Independent Scientific Advisory Committee (ISAC 15_246R). This study did not directly involve patients, and so further ethical approval was not required.

### Data/code availability

This analysis used the CPRD-GOLD primary care dataset March 2016 snapshot (ISAC 15_246R), which is available upon application to the CPRD Independent Scientific Advisory Committee. The code lists used to define the outcomes and covariates for this study are available as a supplementary file. The code lists, as well as the scripts used to create the study cohort and perform the analyses, are also available from the GitHub repository for this project (https://github.com/mcguinlu/CPRD-LRA), which was archived at the time of publication on Zenodo (DOI: **TBC**).

### Supplementary data

Supplementary data are available at IJE online.

