## Supplementary Tables/Figures for "Association of lipid-regulating drugs with dementia and related conditions: an observational study of data from the Clinical Practice Research Datalink"

Supplementary Files updated: 13 October, 2021

### 1 Supplementary Materials

#### 1.1 Supplementary Material 1: Changes to protocol

Any changes made to the approved protocol are “Minor amendments” as per the ISAC criteria, specifically falling under the category of “Additional methods to further control for confounding or sensitivity analysis provided these are to be reported as secondary to the main findings.”

Main changes include:

- Use of a time-varying treatment indicator, to correctly classify time-at-risk.
- Inclusion of additional covariates that are adjusted for in the main model.

#### 1.2 Supplementary Material 2: RECORD Reporting Guidelines

A copy of the RECORD Checklist(1) is provided below.

| **#** | **STROBE items** | **RECORD items** | **Location in manuscript where items are reported** |
| --- | --- | --- | --- |
| 1 | (a) Indicate the study’s design with a commonly used term in the title or the abstract (b) Provide in the abstract an informative and balanced summary of what was done and what was found | RECORD 1.1: The type of data used should be specified in the title or abstract. When possible, the name of the databases used should be included.  RECORD 1.2: If applicable, the geographic region and timeframe within which the study took place should be reported in the title or abstract.  RECORD 1.3: If linkage between databases was conducted for the study, this should be clearly stated in the title or abstract. | *Abstract* |
| 2 | Explain the scientific background and rationale for the investigation being reported |  | *Introduction* |
| 3 | State specific objectives, including any prespecified hypotheses |  | *Introduction* |
| 4 | Present key elements of study design early in the paper |  | *Methods -Section 5.1* |
| 5 | Describe the setting, locations, and relevant dates, including periods of recruitment, exposure, follow-up, and data collection |  | *Methods -Section 5.1/5.2* |
| 6 | *(a) Cohort study* - Give the eligibility criteria, and the sources and methods of selection of participants. Describe methods of follow-up  *Case-control study* - Give the eligibility criteria, and the sources and methods of case ascertainment and control selection. Give the rationale for the choice of cases and controls  *Cross-sectional study* - Give the eligibility criteria, and the sources and methods of selection of participants  *(b) Cohort study* - For matched studies, give matching criteria and number of exposed and unexposed  *Case-control study* - For matched studies, give matching criteria and the number of controls per case | RECORD 6.1: The methods of study population selection (such as codes or algorithms used to identify subjects) should be listed in detail. If this is not possible, an explanation should be provided.  RECORD 6.2: Any validation studies of the codes or algorithms used to select the population should be referenced. If validation was conducted for this study and not published elsewhere, detailed methods and results should be provided.  RECORD 6.3: If the study involved linkage of databases, consider use of a flow diagram or other graphical display to demonstrate the data linkage process, including the number of individuals with linked data at each stage. | *Described in Methods -Section 5.2, and codelists available via linked GitHub repository*  *NA*  *NA* |
| 7 | Clearly define all outcomes, exposures, predictors, potential confounders, and effect modifiers. Give diagnostic criteria, if applicable. | RECORD 7.1: A complete list of codes and algorithms used to classify exposures, outcomes, confounders, and effect modifiers should be provided. If these cannot be reported, an explanation should be provided. | *Available via linked GitHub repository* |
| 8 | For each variable of interest, give sources of data and details of methods of assessment (measurement).  Describe comparability of assessment methods if there is more than one group |  | *Methods -Sections 5.2-5.4* |
| 9 | Describe any efforts to address potential sources of bias |  | *Methods -Section 5.7* |
| 10 | Explain how the study size was arrived at |  | *Methods -Section 5.1* |
| 11 | Explain how quantitative variables were handled in the analyses. If applicable, describe which groupings were chosen, and why |  | *Methods -Sections 5.2-5.5* |
| 12 | (a) Describe all statistical methods, including those used to control for confounding  (b) Describe any methods used to examine subgroups and interactions  (c) Explain how missing data were addressed  (d) *Cohort study* - If applicable, explain how loss to follow-up was addressed  *Case-control study* - If applicable, explain how matching of cases and controls was addressed  *Cross-sectional study* - If applicable, describe analytical methods taking account of sampling strategy  (e) Describe any sensitivity analyses |  | *Methods -Section 5.5*  *Methods -Section 5.7*  *Methods -Section 5.6*  *NA*  *Methods -Section 5.7* |
|  | .. | RECORD 12.1: Authors should describe the extent to which the investigators had access to the database population used to create the study population.  RECORD 12.2: Authors should provide information on the data cleaning methods used in the study. | *Methods -Section 5.1*  *Cleaning script available via linked GitHub repository* |
|  | .. | RECORD 12.3: State whether the study included person-level, institutional-level, or other data linkage across two or more databases. The methods of linkage and methods of linkage quality evaluation should be provided. | *NA* |
| 13 | (a) Report the numbers of individuals at each stage of the study (*e.g.*, numbers potentially eligible, examined for eligibility, confirmed eligible, included in the study, completing follow-up, and analysed)  (b) Give reasons for non-participation at each stage.  (c) Consider use of a flow diagram | RECORD 13.1: Describe in detail the selection of the persons included in the study (*i.e.,* study population selection) including filtering based on data quality, data availability and linkage. The selection of included persons can be described in the text and/or by means of the study flow diagram. | *Supplementary Figure 2* |
| 14 | (a) Give characteristics of study participants (*e.g.*, demographic, clinical, social) and information on exposures and potential confounders  (b) Indicate the number of participants with missing data for each variable of interest  (c) *Cohort study* - summarise follow-up time (*e.g.*, average and total amount) |  | *Methods -Section 6.1 & 6.2, Tables 1 & 2* |
| 15 | *Cohort study* - Report numbers of outcome events or summary measures over time  *Case-control study* - Report numbers in each exposure category, or summary measures of exposure  *Cross-sectional study* - Report numbers of outcome events or summary measures |  | *Table 1* |
| 16 | (a) Give unadjusted estimates and, if applicable, confounder-adjusted estimates and their precision (e.g., 95% confidence interval). Make clear which confounders were adjusted for and why they were included  (b) Report category boundaries when continuous variables were categorized  (c) If relevant, consider translating estimates of relative risk into absolute risk for a meaningful time period |  | *Supplementary Figure 3*  *Methods – Section 5.5*  *NA* |
| 17 | Report other analyses done—e.g., analyses of subgroups and interactions, and sensitivity analyses |  | *Results - Section 6.4* |
| 18 | Summarise key results with reference to study objectives |  | *Discussion - Section 7.1* |
| 19 | Discuss limitations of the study, taking into account sources of potential bias or imprecision. Discuss both direction and magnitude of any potential bias | RECORD 19.1: Discuss the implications of using data that were not created or collected to answer the specific research question(s). Include discussion of misclassification bias, unmeasured confounding, missing data, and changing eligibility over time, as they pertain to the study being reported. | *Discussion - Section 7.3* |
| 20 | Give a cautious overall interpretation of results considering objectives, limitations, multiplicity of analyses, results from similar studies, and other relevant evidence |  | *Conclusions* |
| 21 | Discuss the generalisability (external validity) of the study results |  | *Conclusions* |
| 22 | Give the source of funding and the role of the funders for the present study and, if applicable, for the original study on which the present article is based |  | *Declarations, Back matter* |
|  | .. | RECORD 22.1: Authors should provide information on how to access any supplemental information such as the study protocol, raw data, or programming code. | *Data/code availability section, Back matter* |

### 2 Supplementary tables

#### 2.1 Supplementary Table 1: Definition of exposures and covariates

The code lists used to define covariates adjusted for in the fully-adjusted model (see Supplementary Table 1) were originally created for use in a previous analysis.(2) Some code lists were built on or adapted from previous published work,(3–5) and these are noted in the table.

Table 1: Definition of covariates adjusted for in the Cox PR model.

| **Covariate** | **How was the covariate defined?** |
| --- | --- |
| **Previous history of coronary arterial disease** | Presence of one or more relevant Read codes on record. |
| **Previous history of coronary bypass surgery** | Presence of one or more relevant Read codes on record. |
| **Previous history of cerebrovascular disease (including stroke)** | Presence of one or more relevant Read codes on record. |
| **Chronic illness, including cancer and arthritis** | Charlson index implemented using Read code lists. (2) Code lists based on those by Taylor et al. (3) |
| **Socioeconomic position** | 2010 English Index of Multiple Deprivation (IMD) at the twentile level, where 1 represents the least deprived and 20 the most deprived. |
| **Consultation rate** | Calculated by dividing the total number of clinic visits by the length of the patient record prior to the index date to give an average annual rate. |
| **Alcohol status** | Recorded value (current, former or never). |
| **Smoking status** | Most recent of recorded value (current, former or never) or Read code indicating a recorded value. Code lists based on those by Wright et al. (4) |
| **Body Mass Index** | Recorded value if available, or a calculated value using the last recorded height and weight measurements. Measurements taken before the age of 25 were excluded to ensure adult measurements were used. |
| **Peripheral arterial disease** | Presence of one or more relevant Read codes on record. |
| **Hypertension** | Presence of one or more relevant Read codes on record. |
| **Baseline total cholesterol** | Continuous value recorded as test result ("enttype==163 & test_data1==3") |
| **Baseline LDL cholesterol** | Continuous value recorded as test result ("enttype==177 & test_data1==3") |
| **Chronic kidney disease** | Presence of one or more relevant Read codes on record. |
| **Type 1 Diabetes** | Presence of one or more relevant Read codes on record. |
| **Type 2 Diabetes** | Presence of one or more relevant Read codes on record. |

#### 2.2 Supplementary Table 2: Adherence and switching by drug class

Table 2: Adherence and switching by drug class.

|  | **Whole Sample** | **Statins** | **Bile acid sequestrants** | **Ezetimibe** | **Ezetimibe & Statins** | **Fibrates** | **Nicotinic acid groups** | **Omega-3 Fatty Acid Groups** |
| --- | --- | --- | --- | --- | --- | --- | --- | --- |
| **Stopped** | 6.9% (115899) | 19.1% (111798) | 56.1% (3028) | 19.7% (150) | 12.6% (16) | 12.3% (478) | 44.8% (74) | 35.8% (355) |
| **Added** | 1.6% (27441) | 4.4% (25990) | 3.6% (192) | 19.0% (145) | 3.9% (5) | 21.6% (841) | 3.6% (6) | 26.4% (262) |
| **Switched** | 0.9% (14935) | 2.0% (11996) | 11.3% (612) | 34.6% (264) | 64.6% (82) | 44.0% (1713) | 45.5% (75) | 19.5% (193) |

### 3 Supplementary figures

#### 3.1 Supplementary Figure 1

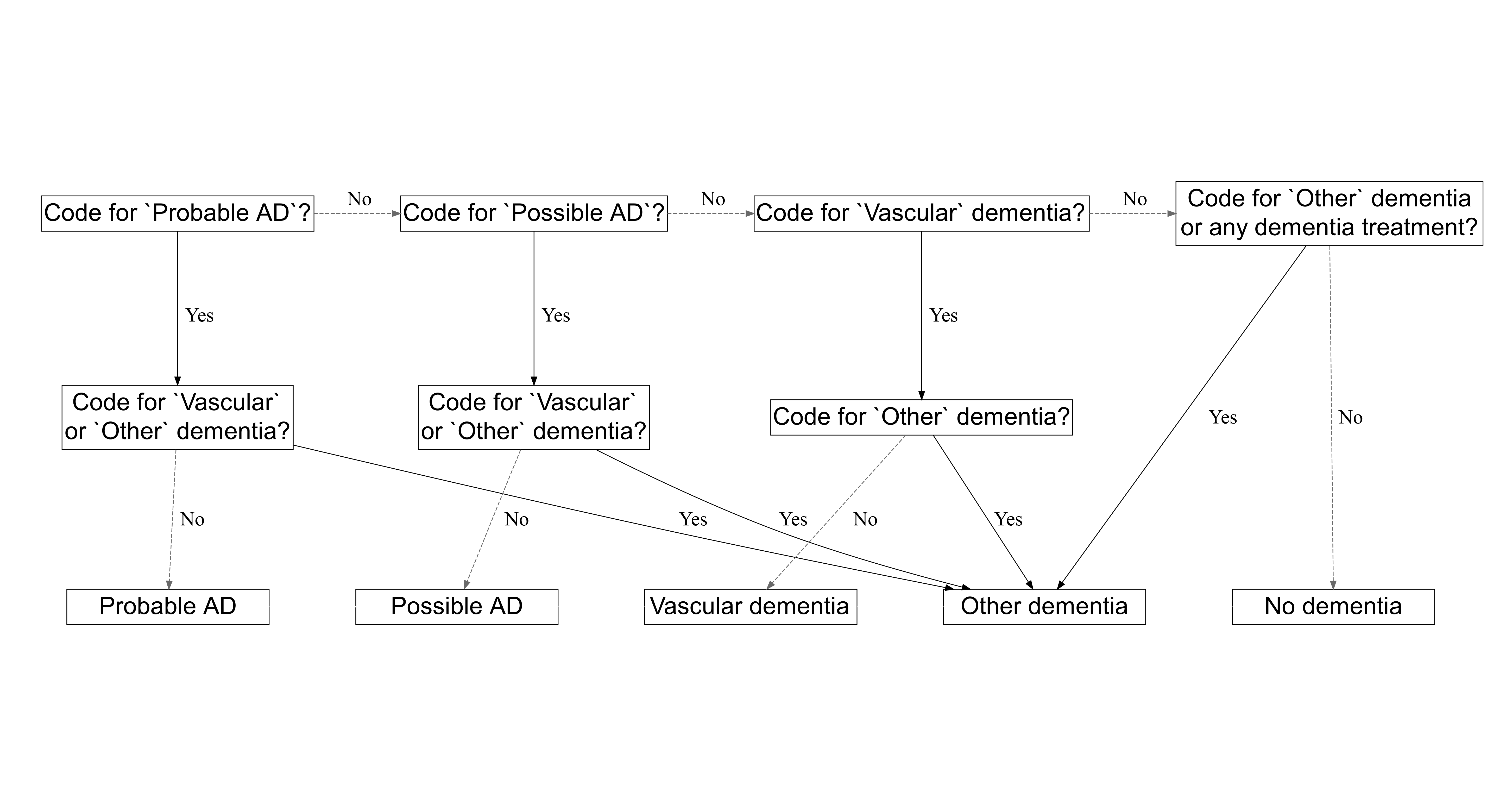

Figure 1: The algorithm used to choose between two diagnosis. This decision tree is adapted with permission from Walker et al (2020).(2)

#### 3.2 Supplementary Figure 2

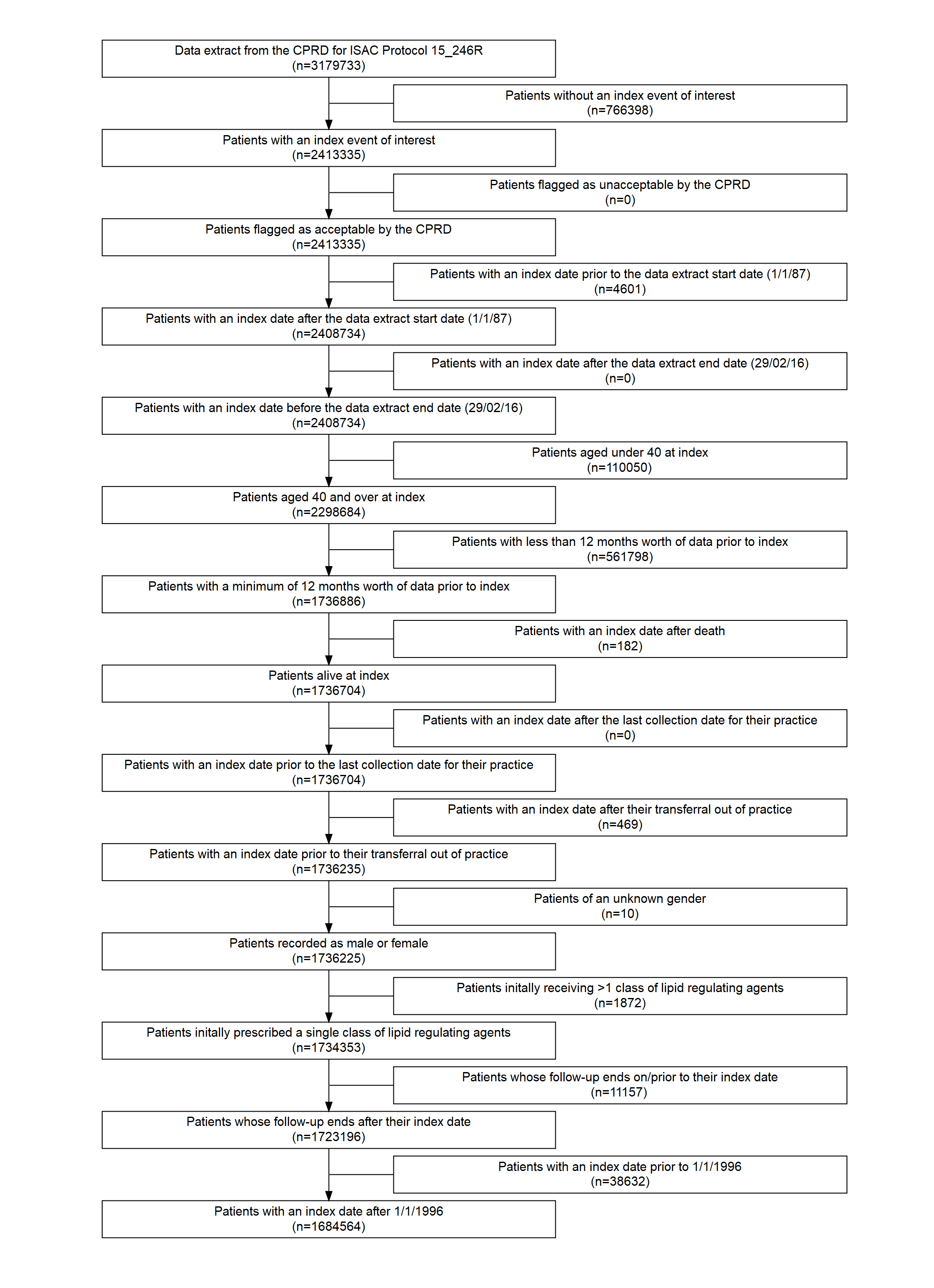

Figure 2: Attrition of participants as the eligibility criteria were applied.

#### 3.3 Supplementary Figure 3

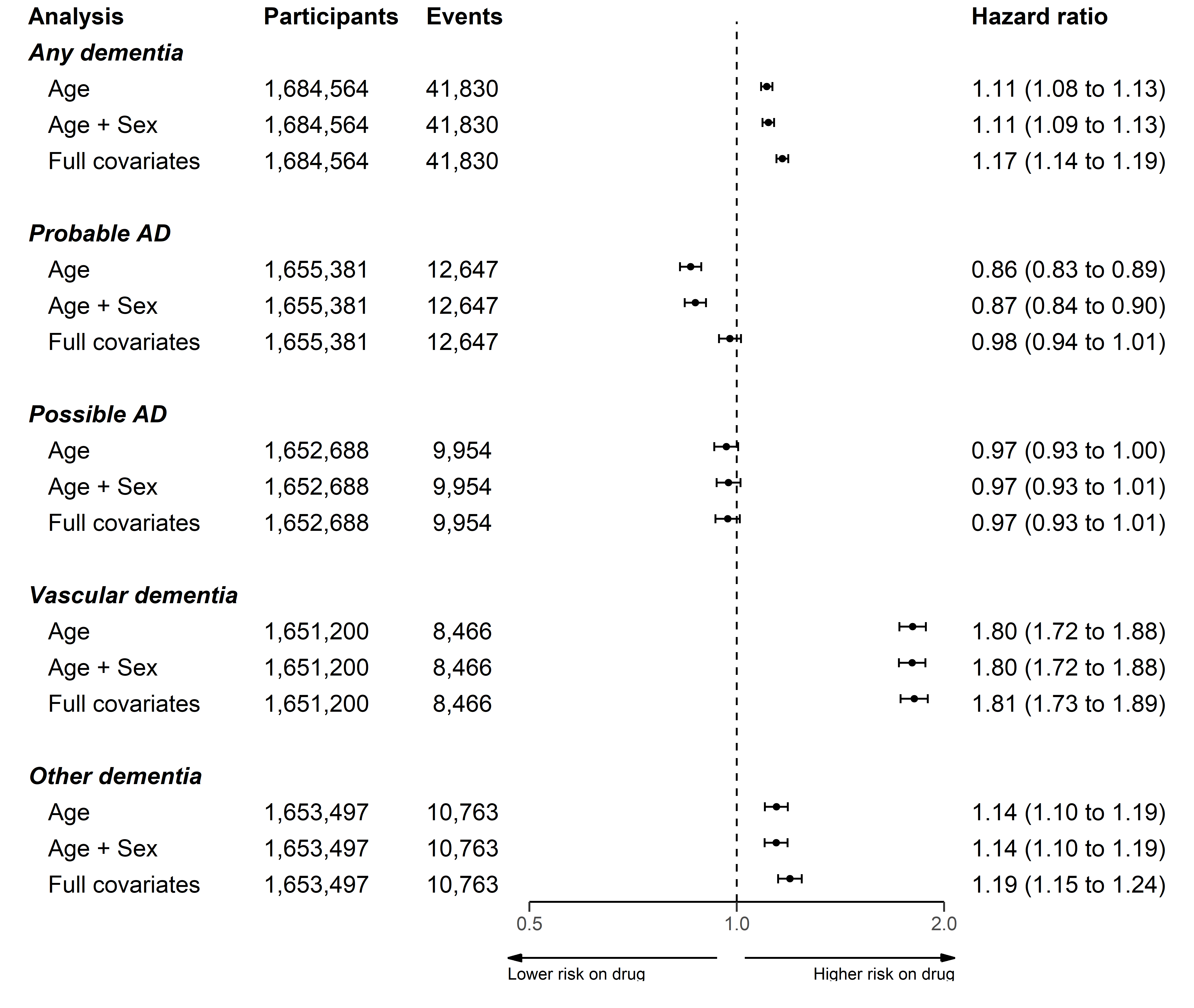

Figure 3: Association of any lipid regulating agent with a dementia or related outcome using three models adjusted for age, age and sex, and all covariates respectively.

#### 3.4 Supplementary Figure 4

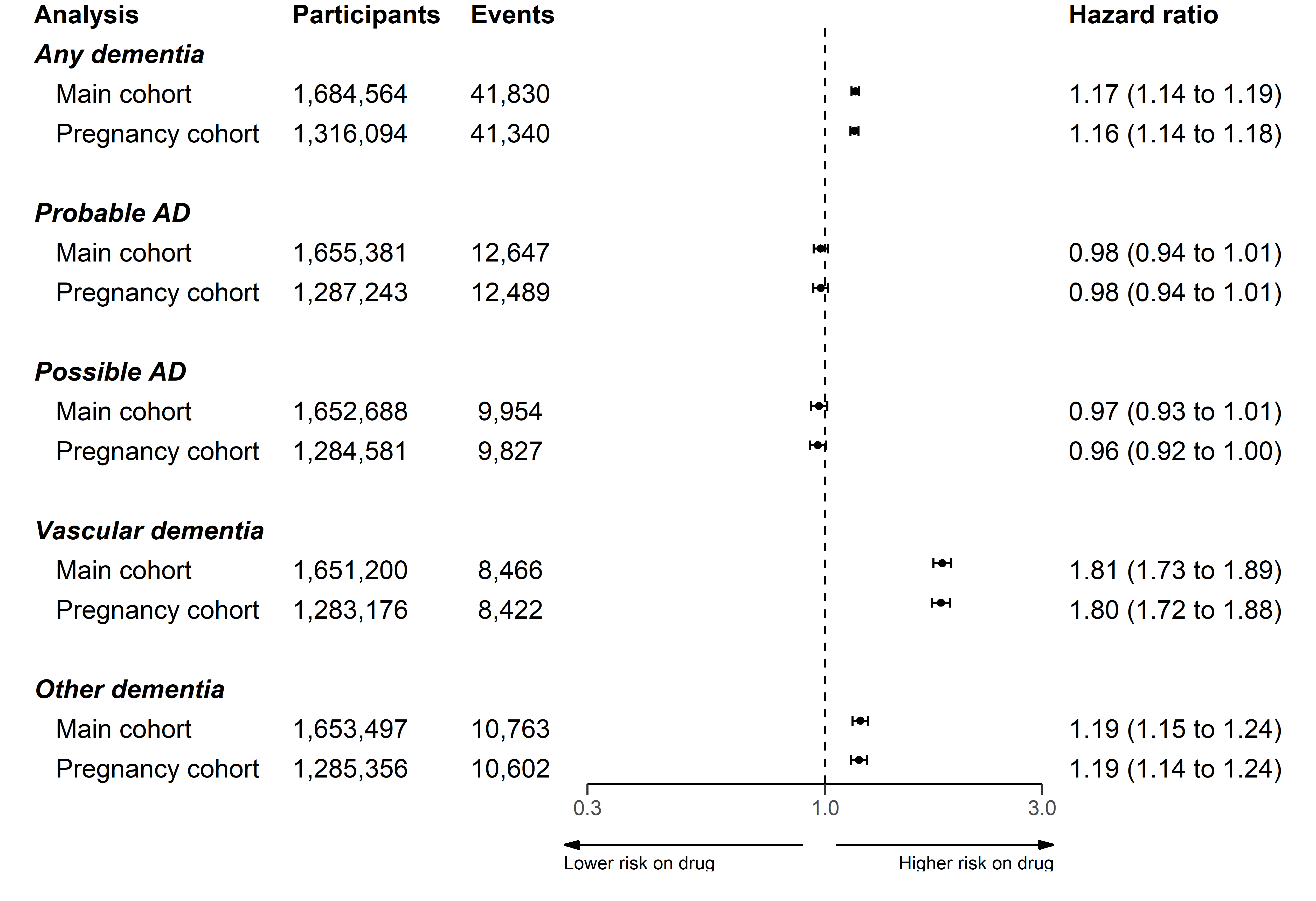

Figure 4: Association of any lipid regulating agent with a dementia or related outcome, removing participants who were less than 55 years of age at index.

#### 3.5 Supplementary Figure 5

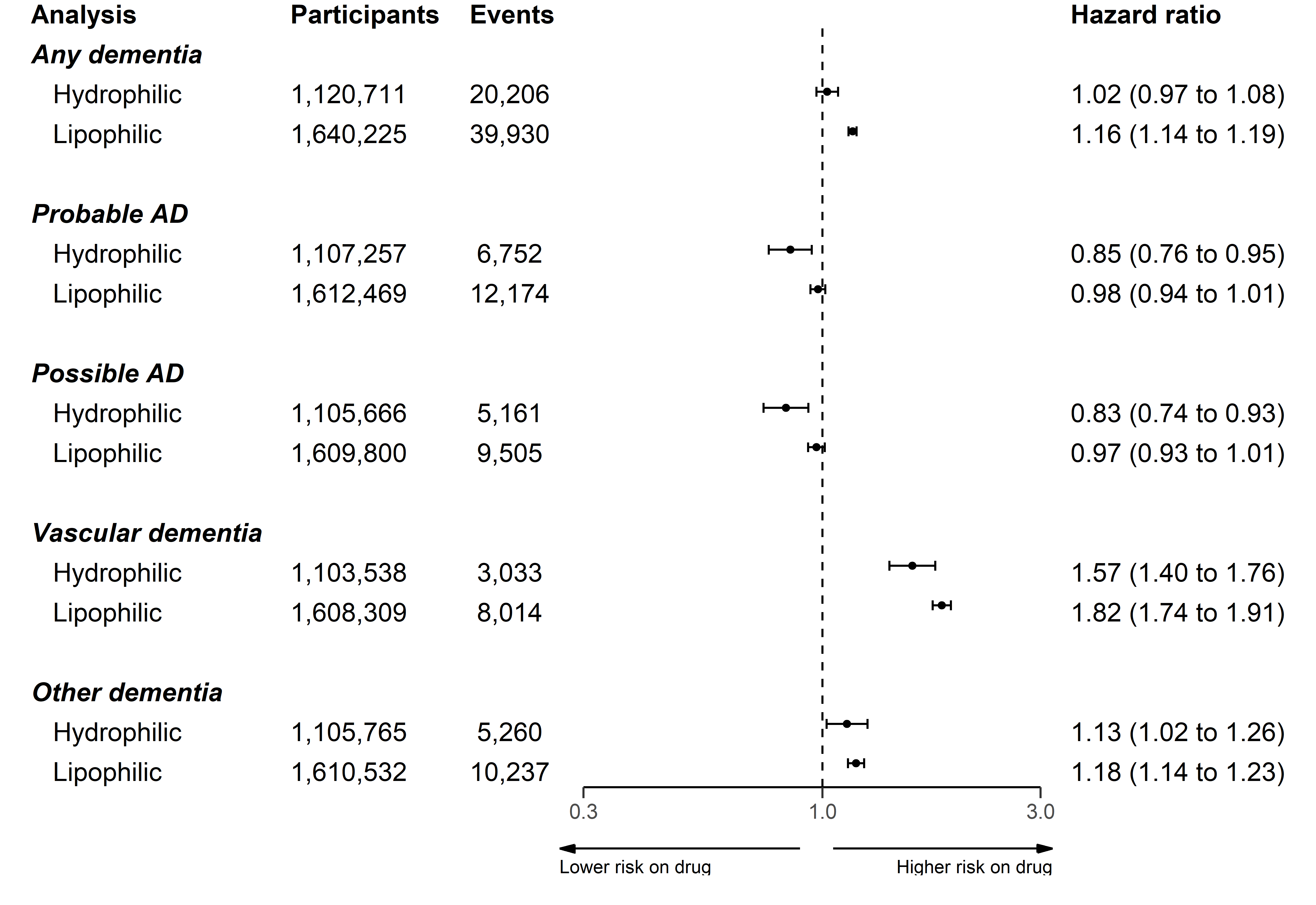

Figure 5: Association of statins with a dementia or related outcome, stratified by statin lipophilic/hydrophilic properties.

#### 3.6 Supplementary Figure 6

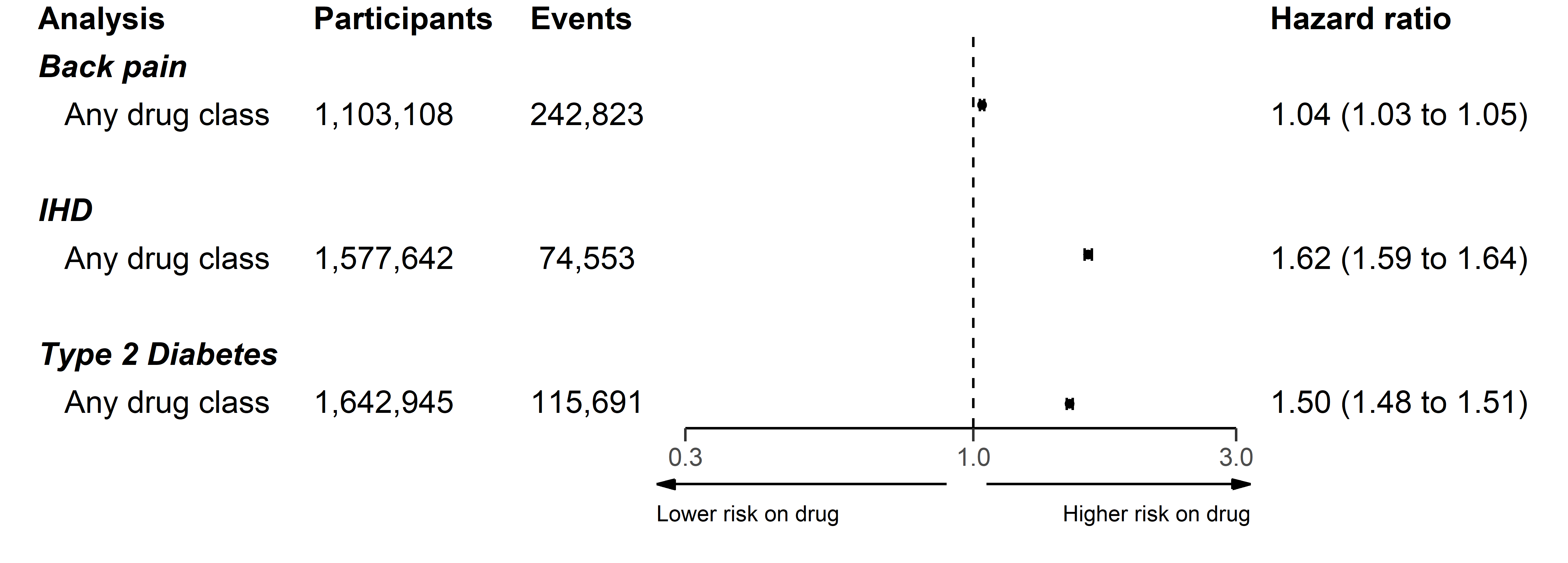

Figure 6: Association of any lipid regulating agent with backpain, ischemic heart disease (IHD), and Type 2 diabetes.
